## Supplementary figures and images for "Characterization of p53 p.T253I as a pathogenic mutation underlying Li-Fraumeni Syndrome"

### Supplemental Figure 1

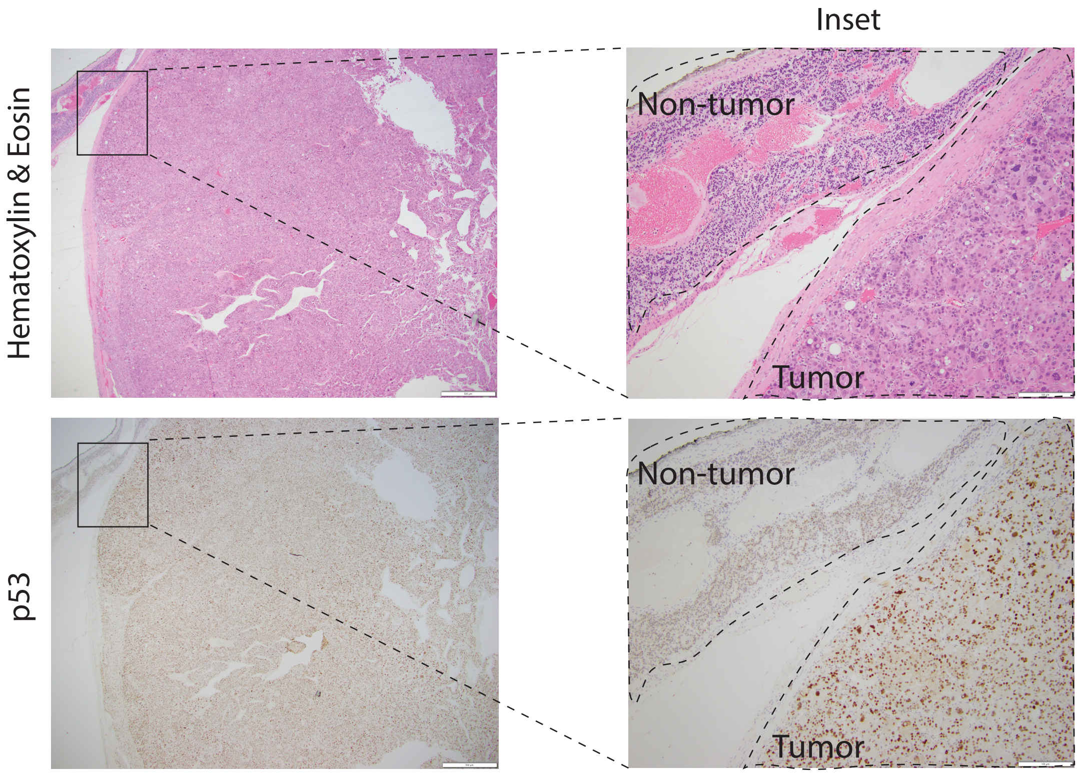

### Supplemental Figure 2

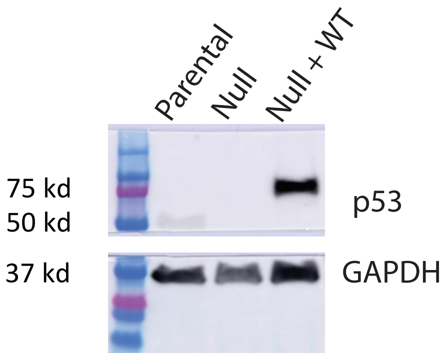

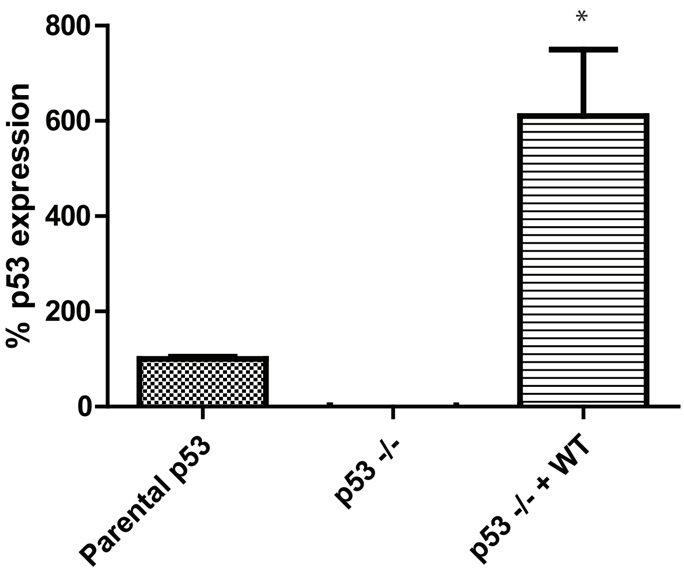

### Supplemental Figure 3

*Holcomb, et. al., Sup. Fig. 3*

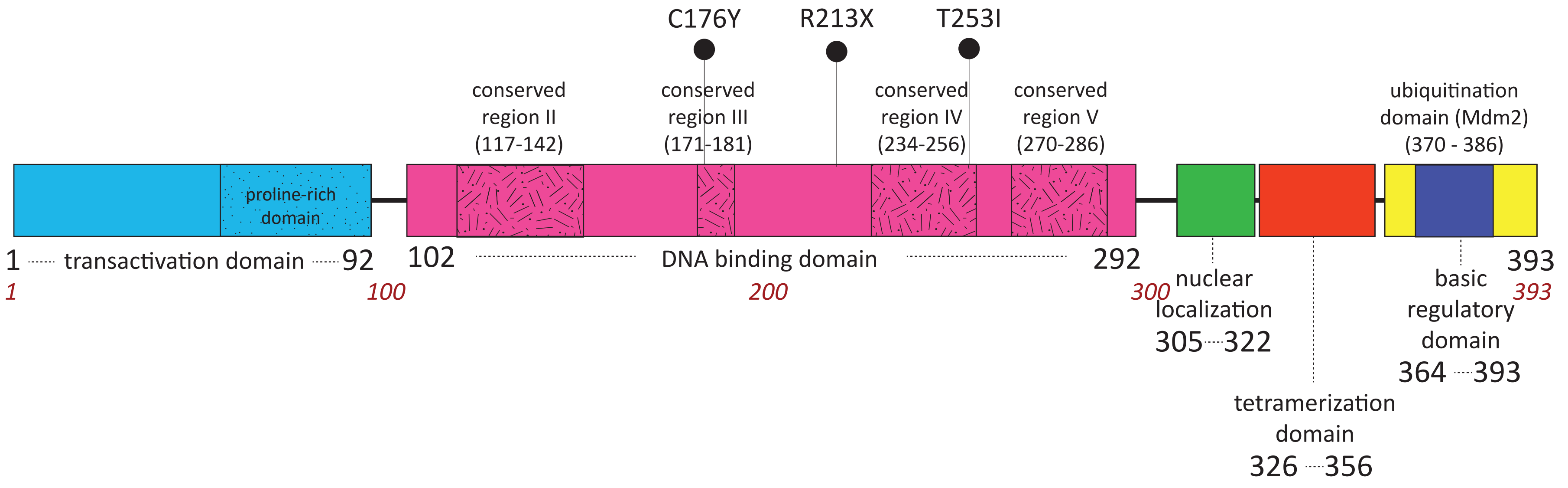

### Supplemental Figure 4

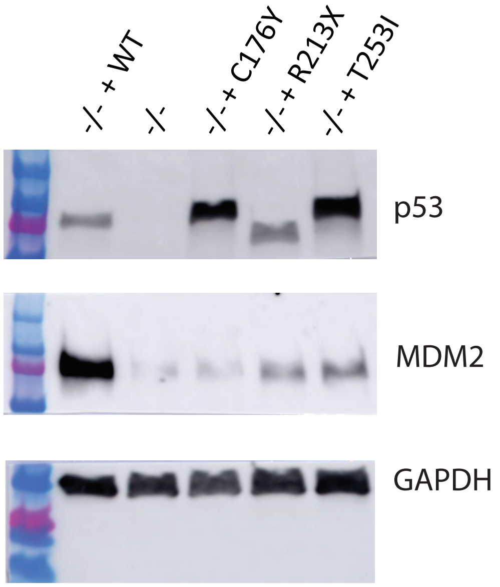

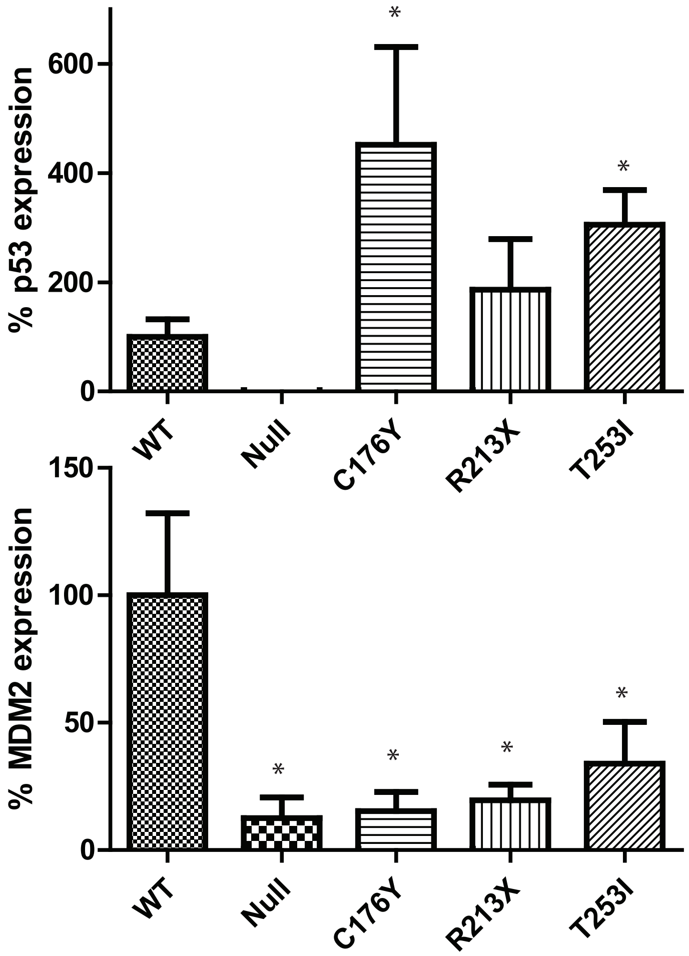

### Supplemental Figure 5

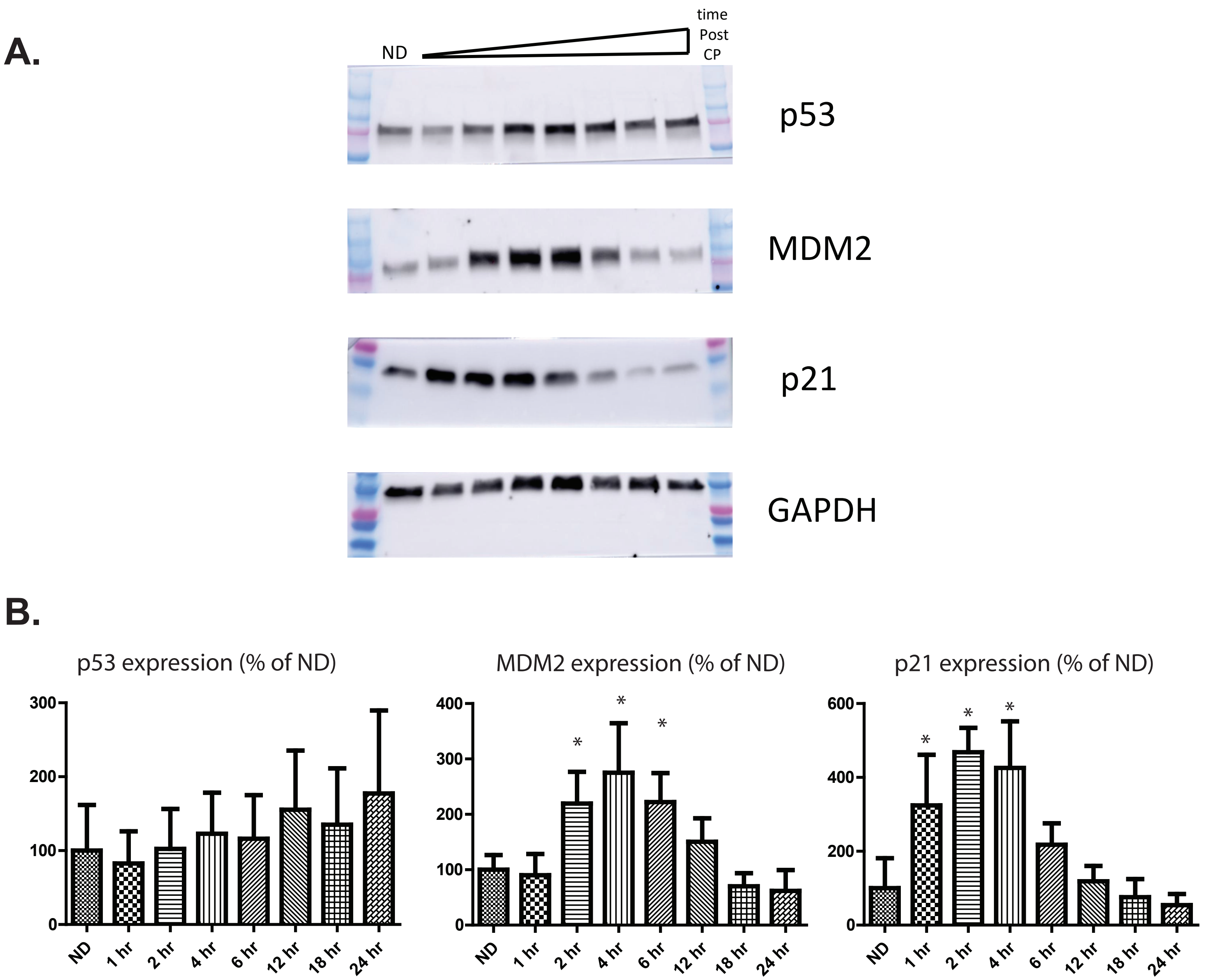

### Supplemental Figure 6

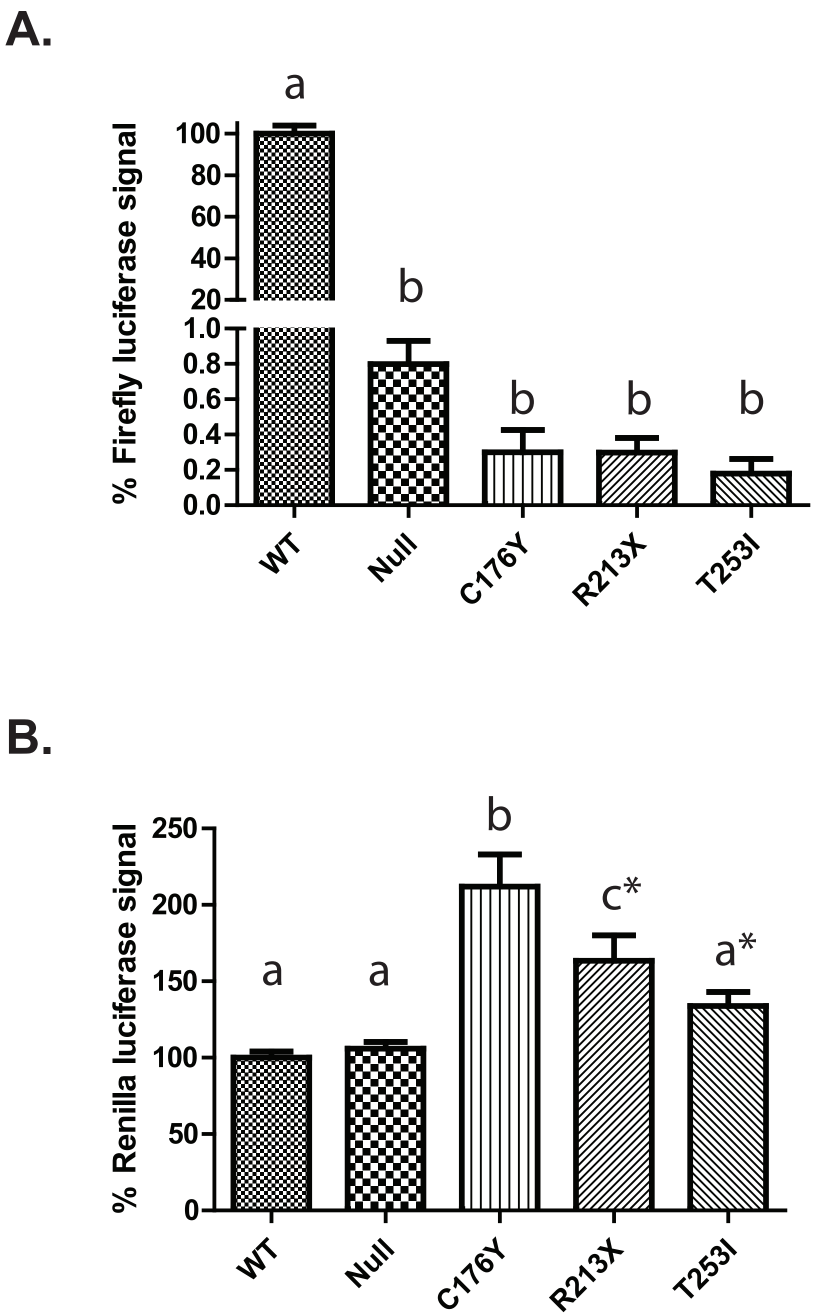
